# Seizure forecasting by tracking cortical response to electrical stimulation

**DOI:** 10.1101/2023.11.25.23298484

**Authors:** Petroula Laiou, Zeljko Kraljevic, Antonio Valentin, Sharon Jewell, Pedro F. Viana, Chirag Mehra, Richard J.B. Dobson, Andreas Schulze-Bonhage, Matthias Dümpelmann, Timothy J. Denison, Joel S. Winston, Mark P. Richardson

## Abstract

**Background:** Seizure unpredictability is a significant burden in the lives of people with epilepsy. Previously published approaches to seizure forecasting analysed intracranial electroencephalographic recordings (iEEG) and showed that seizures can be forecast above chance levels. Although passive observation of the brain might provide some insights, repeated active perturbation of the cortex and measuring the cortical response may provide more direct information about time-varying cortical excitability.

**Objective:** The aim of this study is to investigate whether seizures can be forecast by stimulating the cortex via intracranial electrodes and measuring cortical response from the iEEG.

**Methods:** We studied a cohort of eight patients with treatment-resistant epilepsy who were admitted to King’s College Hospital for presurgical evaluation with iEEG. During their stay, they underwent prolonged single pulse electrical stimulation for approximately one day. Stimuli were delivered every 5 minutes to a constant pair of electrodes and all patients experienced at least one clinical seizure during the period of stimulation. We extracted quantitative features from the iEEG post-stimulus response and developed a logistic regression algorithm to estimate the seizure likelihood at each stimulus. To evaluate the algorithm’s performance, we used improvement over chance (IoC), sensitivity, time spent in warning and Brier Skill score. We also compared performance with seizure prediction based on passive observation of iEEG.

**Results:** In seven out of eight patients, seizures could be forecast using the post-stimulus response above chance levels (average IoC: 0.74). In comparison, the seizure forecasting performance based on passive (unstimulated) iEEG was less good (average IoC: 0.54).

**Conclusions:** These results suggest that cortical response to electrical stimulation may aid in the development of seizure forecasting algorithms as well as in the design of novel implantable devices that deliver electrical stimulation to control seizures.

## Introduction

Globally, around 65 million people have epilepsy, and 5 million new cases are diagnosed per year, making it one of the most common neurological disorders [Milligan_2021, WHO_2023]. The key feature of epilepsy is the occurrence of recurrent seizures, and the first line of treatment is the administration of antiepileptic drugs (AEDs). Although AEDs are able to control seizures in around two thirds of people, the remaining one third suffer from uncontrolled seizures that create a significant burden in their lives [Chen_2018]. The ability to forecast the occurrence of upcoming seizures would significantly increase the quality of life of people with epilepsy by providing a seizure warning system, which could be used to enhance safety and to allow the application of preventative therapy.

Several studies that analysed continuous biosignals such as intracranial electroencephalography (iEEG), scalp EEG, heart rate, electrodermal activity and accelerometer measurements demonstrated that seizures can be forecast above chance levels [Cook_2013, Karoly_2017, Baud_2018, Kuhlman_2018, Meisel_2020, Maturana_2020, Proix_2021, Stirling_2021, Brinkmann_2023]. The most successful approaches to seizure forecasting analysed long-term, passively collected iEEG from implantable devices and made seizure-risk forecasts for various time horizons [Cook_2013, Karoly_2017, Baud_2018, Kuhlman_2018, Maturana_2020, Proix_2021]. The mechanism underlying the time-varying seizure-risk is not known yet but is assumed to relate to time-varying cortical excitability. Therefore, measuring and tracking cortical excitability more directly might provide valuable information about seizure forecasting. Active perturbation of the cortex and measuring its response may provide a more direct way to track cortical excitability.

The first reported study of perturbing the cortex to quantify cortical excitability changes that occur prior to seizures used MEG (magnetoencephalography) and EEG responses to intermittent photic stimulation in people with photosensitive epilepsy [Kalitzin_2002]. This study showed that a phase-based quantitative measure (rPCI) significantly increased prior to seizures. The same group [Kalitzin_2005] subsequently analysed iEEG responses to electrical stimulation and demonstrated that in people with mesial Temporal Lobe Epilepsy (mTLE) elevated rPCI values correlated with shorter time intervals to the next seizure. In another study [Freestone_2011], the temporal evolution of quantitative metrics estimated from the iEEG responses to electrical stimulation was tracked in two patients with TLE and showed variation across the sleep-wake cycle and seizure occurrence. This study showed promise that probing the cortex with electrical stimulation and measuring its response using iEEG could be informative for seizure forecasting.

In the study presented here, we investigated whether we could forecast seizures by tracking cortical response to electrical stimulation. We studied a cohort of eight epilepsy patients that were implanted with intracranial electrodes and underwent prolonged intermittent electrical stimulation for one day. We extracted quantitative features at each stimulus and developed a logistic regression algorithm to estimate the seizure likelihood in each stimulus. In addition, we investigated how the seizure forecasting performance computed from the stimulus response compared to forecasts that were estimated using passive observation of iEEG.

## Methods

### Patients and data collection

We studied a cohort of eight patients with treatment-resistant focal epilepsy (5 male; mean age: 34.8 years) who were admitted to King’s College Hospital for presurgical evaluation using iEEG. All patients were implanted with subdural (strip, grid) or depth electrodes (Ad-Tech Medical Instruments Corp., WI, USA) whose type, number and location were determined by the clinical team for each patient individually. At King’s College Hospital the administration of single pulse electrical stimulation (SPES) is part of the presurgical protocol. The SPES protocol has been described in detail in Valentin et al. [Valentin_2002, Valentin_2005]. In brief, 10 single monophasic pulses (1ms duration; current intensity 2-5mA) with a gap of 10 sec are systematically delivered to all neighbouring electrodes to map cortical excitability. SPES provoke two main types of cortical responses: the early and late response. Early cortical responses to SPES that occur within 100ms after stimulation are considered physiological responses of the cortex to SPES. Late cortical responses to SPES that occur between 100ms-1sec after stimuli are considered abnormal [Valentin_2002]. It has been shown that the surgical removal of the brain tissue that corresponds to regions that generate late SPES responses is associated with the seizure focus and good-postsurgical outcome [Valentin_2005].

Once the clinical team collected all the clinical information from the video-iEEG monitoring, and before the explantation of the iEEG electrodes, the patients underwent intermittent stimulation for approximately one day. If antiseizure medications had been reduced or withdrawn, they were restored prior to the period of stimulation. In each patient, the pair of electrodes that generated the most obvious late SPES responses was selected for the prolonged stimulation and two single monophasic pulses (1ms duration; current intensity 2-5mA) that were separated by 5sec were delivered every 5min using a constant current neurostimulator (Medelec ST10 Sensor, Oxford Instruments). This stimulation procedure lasted from 12 to 26 hours (mean: 18 hours; Table 1) and none of the patients reported any behavioural percept. The number of clinical seizures in each patient varied from one to three. In the analysis we only considered the first seizure as the remaining seizures occurred in short time intervals i.e., in less than three hours from the first seizure. The study was approved by the ethics committee of King’s College Hospital (Reference number: 06/Q0703/117) and all patients gave written informed consent to participate in the study.

**Table 1.**
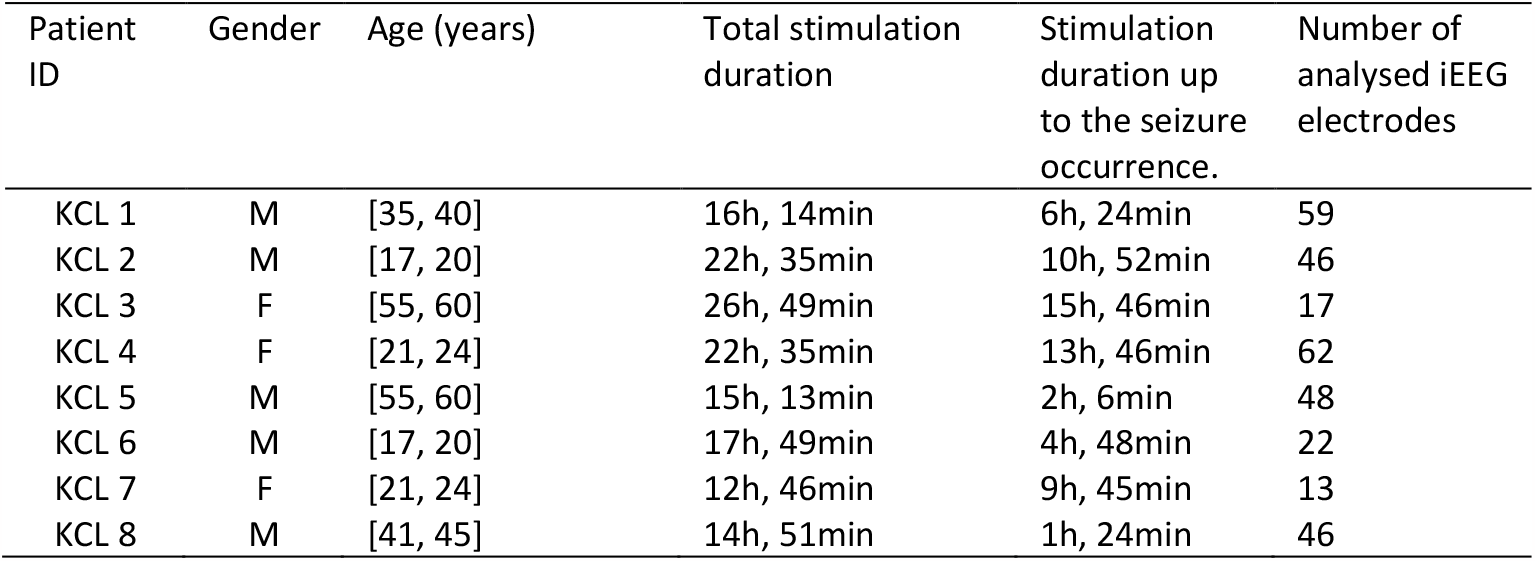
Patient information.

| Patient ID | Gender | Age (years) | Total stimulation duration | Stimulation duration up to the seizure occurrence. | Number of analysed iEEG electrodes |
| --- | --- | --- | --- | --- | --- |
| KCL 1 | M | [35, 40] | 16h, 14min | 6h, 24min | 59 |
| KCL 2 | M | [17, 20] | 22h, 35min | 10h, 52min | 46 |
| KCL 3 | F | [55, 60] | 26h, 49min | 15h, 46min | 17 |
| KCL 4 | F | [21, 24] | 22h, 35min | 13h, 46min | 62 |
| KCL 5 | M | [55, 60] | 15h, 13min | 2h, 6min | 48 |
| KCL 6 | M | [17, 20] | 17h, 49min | 4h, 48min | 22 |
| KCL 7 | F | [21, 24] | 12h, 46min | 9h, 45min | 13 |
| KCL 8 | M | [41, 45] | 14h, 51min | 1h, 24min | 46 |

### Signal Preprocessing

After the iEEG acquisition, all iEEG signals were epoched in 4sec segments that were centred around each stimulus. Next, all iEEG epochs were visually inspected and epochs that had corrupted signals were removed from the analysis. All iEEG signals were re-referenced to the average and downsampled to 256Hz. To eliminate the stimulation artifact, the data points 0-20 ms post-stimulus were removed and replaced using a cubic spline interpolation. Afterwards, all signals were band-pass filtered (forward and backward filtering to minimize phase distortion) between 0.1 and 120 Hz and notch filtered between 48 and 52Hz using a fourth-order Butterworth filter. Figure 1 illustrates a schematic diagram of the analysis steps.

**Figure 1.**
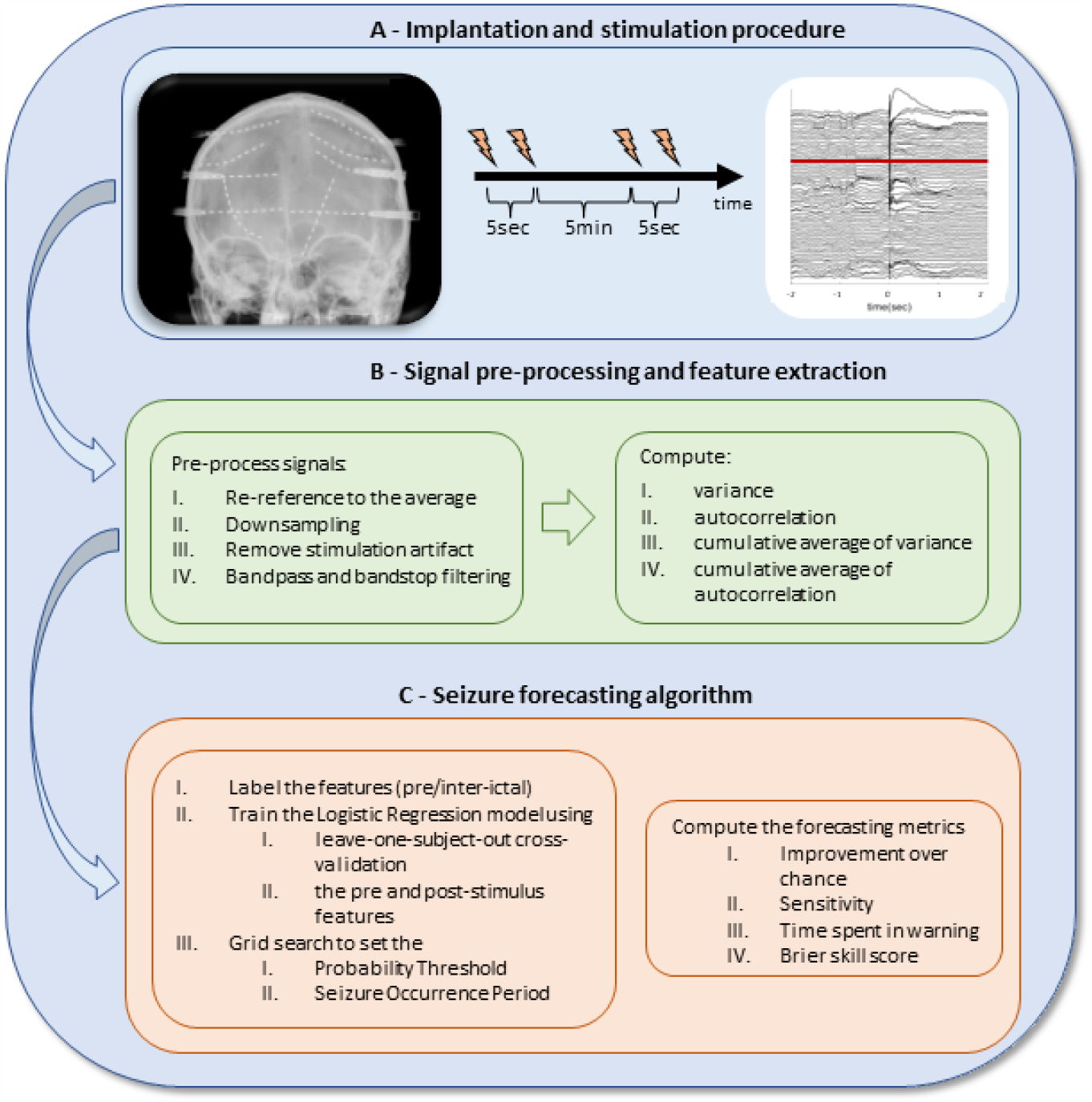
Schematic representation of the study design. Two electrical pulses separated by 5sec were administered to a constant pair of electrodes every 5min (A). After signal pre-processing, quantitative features were extracted from the post and pre-stimulus response (B). Next a Logistic regression classifier was applied using a leave-one-subject-out cross validation approach. Forecasting metrics quantified algorithms performance (C).

### Selection of time windows relative to stimulations

Every five minutes, a pair of stimuli were delivered with a separation of 5 s. We intended to extract features around these pairs of stimuli but had no a priori information about the relevant window timing or duration, therefore we examined several time windows. We examined short windows (20ms to 100ms post the first stimulus and post the second stimulus) and longer windows (20ms to 1000ms post the first stimulus and post the second stimulus). Preliminary analysis of data suggested that there might be long duration effects of stimulation, hence we also examined a window 4000ms to 4980ms after the first stimulus (to leave a 20ms buffer prior to the second stimulus). Finally, we also examined a time window prior to the first stimulus, i.e., -1000ms to -20ms pre-stimulation to obtain information about ‘passive’ unstimulated features.

### Selection of electrodes

Each patient had a different number of electrodes distributed in various anatomical locations. To standardize the approach across patients, we examined data from three different electrode sets: 1) the single iEEG signal that manifested the most prominent response across the stimulation procedure; 2) five iEEG signals that showed the most prominent response across the stimulation (we chose five because many implantable devices that deliver electrical stimulation are usually equipped with two to eight iEEG electrodes and five is the median of this range); and 3) all available iEEG signals. Note that the pair of stimulated signals was excluded from the analysis.

### Feature Extraction

Cortical response to electrical stimulation was quantifying using the variance 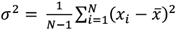, where *N* is the number of samples of the signal *x*, and autocorrelation 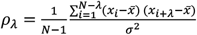 as a function of the lag *λ*. The autocorrelation metric was computed as the width at the half maximum of the autocorrelation function [Maturana_2020]. For the cases for which we considered more than one signal we computed the average of variance and autocorrelation across the analysed signals. For each value of variance and autocorrelation, we also computed its cumulative average across its previous 12 values that correspond to a one hour interval (i.e., 12 × 5 min = 1*hour*). Hence, for a given number of analysed iEEG signals (one, five or all), a given analysis window (one pre-stimulation window, three windows post the first stimulation, and two windows post the second stimulation) we obtained four features, i.e., variance, autocorrelation, cumulative average of variance and cumulative average of autocorrelation.

Features in the 10min time interval before and after each seizure timestamp were removed from the analysis. In addition, all features were smoothed using a backward moving average filter with a 30min window length (i.e., average between the current and previous five points) to eliminate spontaneous fluctuations. Specifically, features were split in two windows that were separated at the timestamp of the seizure. Smoothing was performed in each window separately and features that corresponded to the first 30min of each window were discarded from the analysis to eliminate edge effects.

### Seizure forecasting algorithm

All features from the 3-hour interval prior to seizure occurrence were labelled as “pre-ictal”, whereas all features that were more that 3-hours from the seizure timestamp were labelled as “inter-ictal”. All features were z-normalized in each patient individually, using the mean and standard deviation of the features distribution during the interictal state. Afterwards, a Logistic Regression classifier was applied with a leave-one-patient out cross-validation approach. Specifically, the algorithm was trained in turns using the feature vectors (i.e., variance, autocorrelation, cumulative average of variance and cumulative average of autocorrelation) from the 7 patients and was tested on the feature dataset from the remaining patient. This process yielded for every patient a probability distribution with the seizure likelihood for each feature vector (i.e., probability likelihood that each feature vector has a “pre-ictal” label).

Afterwards, we employed a grid search to set the optimal “probability threshold” of the probability distribution, such that when it is crossed an alarm would be initiated that a seizure is about to happen in the next time-period i.e., the “seizure occurrence period”. Once this “seizure occurrence period” has passed, then a new alarm can be initiated. In the grid search the “probability threshold” values ranged from 0.4 to 0.8 in steps of 0.01, whereas the “seizure occurrence period” values ranged from 30min to 240min, and we only used the training data to ensure that no information of the test participant is used to set these parameters. The parameter combination that gave the optimal improvement over chance was selected for the analysis. Once these parameters are set, the “Forecasting horizon” denotes the time (in minutes) between the alarm onset until the seizure onset.

To quantify the performance of the seizure forecasting algorithm, we used previously described forecasting metrics [Cook_2013]. Specifically, we used Sensitivity (*S* = *TP*/(*TP* + *FN*), where *TP*: seizures that occur when the alarm is on, *FN*: seizures that occur outside alarms), time spent in warning (*tiw*: the proportion of time that was spent in warning computed as the total number of points for which the alarm was on over the total number of points), Improvement over chance (*IoC* = *Sensitivity − tiw*) and Brier Skill Score that quantifies the improvement of the Brier score relative to a random reference (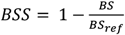 where BS is the Brier Score 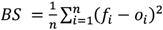, *n* is the number of forecasted points, *fi* forecasted probability of the *i*^*t*h^ forecasted point, *oi* observed value of the *i*^*t*h^ point (0 when the point had “inter-ictal” label and 1 when it had the “pre-ictal” label). *BS*_*ref*_ was computed by randomly shuffling the probability forecasts 100 times and afterwards taking the average BSS value). When there is not improvement over reference then BSS tends to 0, when it is worse than reference it tends to *−* ∞ and to 1 when it is perfect. In addition, to show that the forecasted metric values are not due to chance, we randomly shuffled the predicted forecasted probabilities 100 times and computed the average sensitivity, tiw and IoC across the 100 runs. The signal pre-processing analysis as well as the feature extraction was performed in MATLAB (MathWorks R2020b), whereas the Logistic Regression analysis was executed in Python (version 3.8.16).

## Results

### Temporal evolution of feature profiles

We observed that there was high variability on the temporal evolution of both variance and autocorrelation regardless the time of the day, the pulse sequence (first or second pulse) as well as the length of time interval i.e., 100ms (Figure S1) or 1000ms (Figure 2A). Additionally, in individual cases there was a prominent increase or decrease in the features prior to seizures.

**Figure 2:**
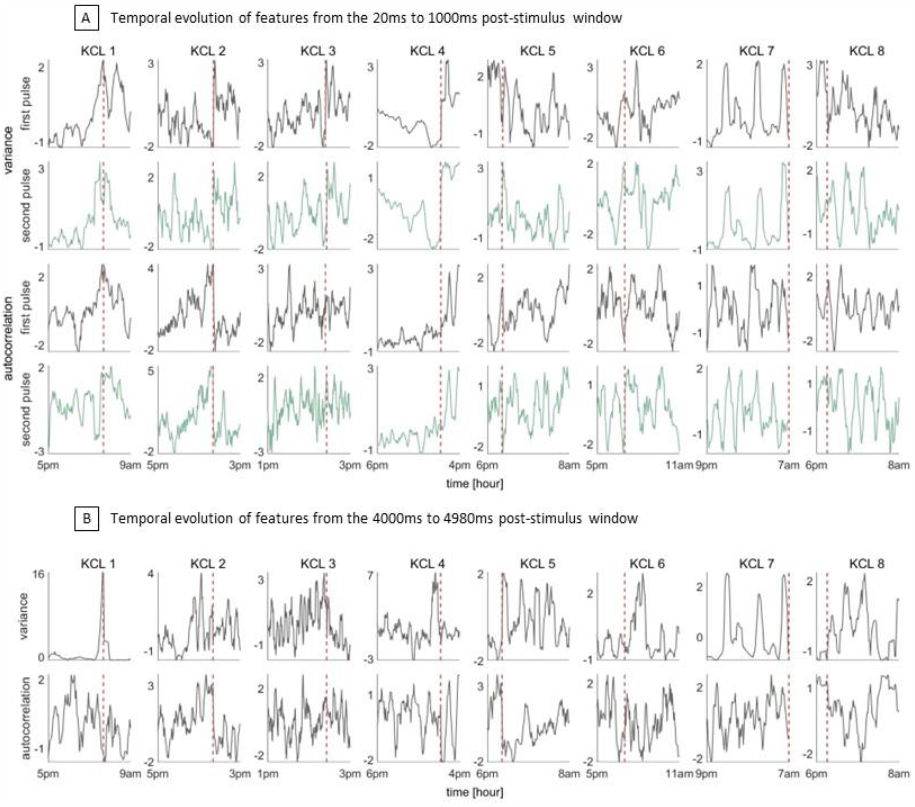
(A) Temporal evolution of the features from the window 20ms to 1000ms after the first stimulus (grey) and after the second stimulus (green). (B) Temporal evolution of the features from the window 4000ms to 4980ms after the first stimulus. The vertical dashed red line denotes the seizure onset.

The temporal evolution of the features in the 4000ms to 4980ms window after the first stimulus is depicted in Figure 2B. We observe that in four patients (KCL1 to KCL4) the profile of the variance shows a prominent increase prior to seizures, whilst in the other four patients the profile of autocorrelation tends to increase prior to seizure occurrence. In addition, the increase in the feature profiles prior to seizures is much clearer and consistent compared to the profiles computed from the 20ms to 100ms post-stimulus window and the 20ms to 1000ms post-stimulus window (Figure S1, Figure 2A).

### Seizure forecasting from the post-stimulus features

Having observed the temporal evolution of the features profiles computed from the various post-stimulus windows, we trained a Logistic Regression (LR) classifier using a leave-one-subject out cross validation approach. A LR classifier was trained using features computed from the 20ms to 100ms post-stimulus window and the 20ms to 1000ms post-stimulus windows, separately for the first and second stimuli (time windows up to 100ms and 1000ms after stimulus). A LR classifier was also trained using features computed from the time interval from 4000ms to 4980ms after the first stimulus. In each case, the output of the LR classifier was a probability distribution with the seizure likelihood at each stimulus.

When we executed the LR classifier using the short-term features computed from the 20ms to 100ms post-stimulus windows from the first or second pulse, the forecasting performance was poor and yielded zero IoC for both pulses (Figure S2A, S2B). The same results hold when we considered the 20ms to 1000ms post-stimulus window of the first pulse (Figure S2C). Interestingly, when the features were computed from the 20ms to 1000ms post-stimulus window of the second pulse, in two out of eight patients we obtained IoC 0.92 and 0.9 (Figure S2D). In contrast, when we analysed features computed from the time interval from 4000ms to 4980ms after the first stimulus, there was a clear increase in the seizure likelihood prior to seizure occurrence in all but one patient (Figure 3A). Across all patients, the average IoC was 0.74, average sensitivity 0.88, average tiw 0.14, average BSS 0.33 and average forecasting horizon 73.86min (Figure 3B). The exact values of the forecasting metrics are provided in Table S1.

**Figure 3:**
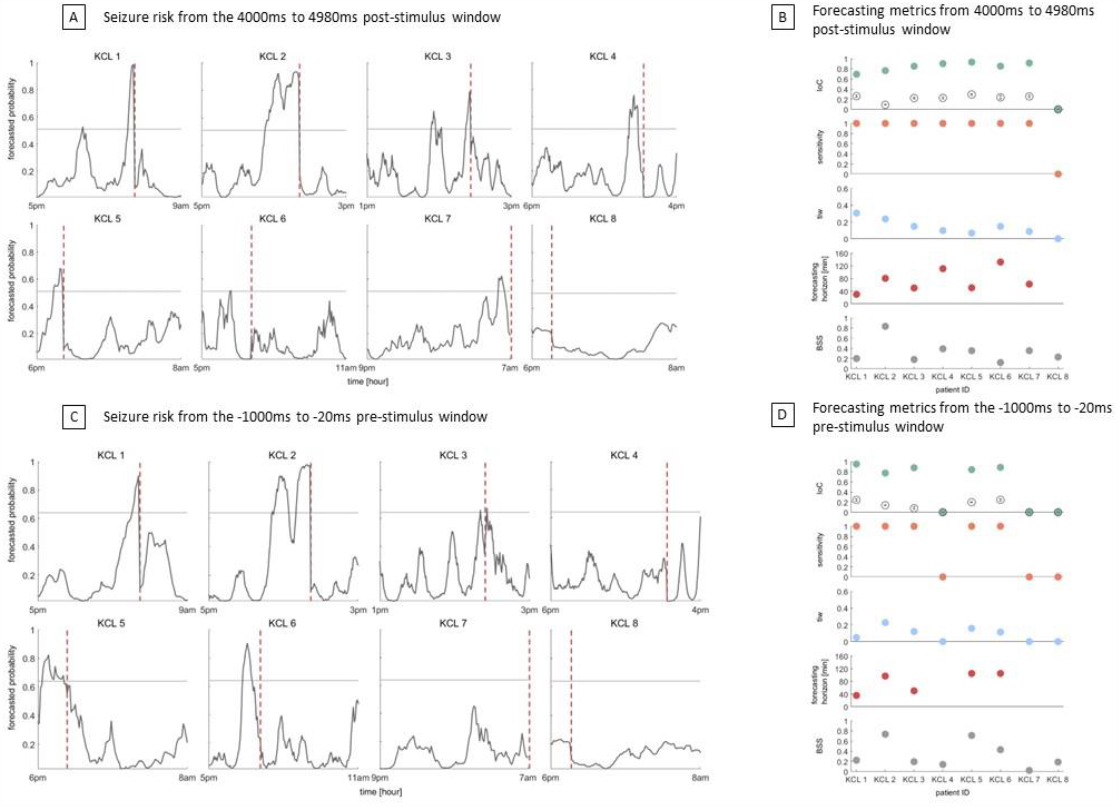
Seizure likelihood and forecasting metrics computed from the window 4000ms to 4980ms after the first stimulus (A, B) and the -1000ms to -20ms window prior to the first stimulus (C, D). The vertical dashed line denotes the seizure onset. After grid search (see Methods) the probability threshold (horizontal line) and seizure occurrence period were set to 0.51; 150min (A) and 0.64; 120min(C) respectively. Grey unfilled circles in panels B and D denote the average IoC obtained from the shuffled forecasts across 100 runs, whilst error bars denote the standard error. Note that negative IoC values from the shuffled forecasts were set to zero prior to averaging.

### Seizure forecasting from the pre-stimulus features

We sought to investigate whether the forecasts from post-stimulation data outperform the forecasts that are computed from passive unstimulated iEEG data. We thus computed the same features from -1000ms to -20ms window prior to the administration of the first stimulus and applied the LR classifier. Note that the time gap from the first pulse and its previous pulse was 5min and hence any effects of stimulation on the cortical excitability would have diminished. Figure 3C illustrates the seizure likelihood for each patient as computed from the LR classifier using the features from the pre-stimulus intervals of the first pulse. In all but three patients (KCL 4, KCL 7, KCL 8) there is an increase in the seizure likelihood before the seizure occurrence. Across all patients the average IoC was 0.54, the average tiw was 0.08, the average BSS was 0.33 and the average forecasting horizon was 78.4min (Figure 3D). The exact values of the forecasting metrics are given in Table S2.

### Mimicking a real-time seizure forecasting system

In the analysis that we performed so far, the feature vectors of each patient were z-normalized using the corresponding mean and standard deviation of all interictal data (see Methods). Hence, the features of the test patient were normalized at each stimulus using future information. In a real-time seizure forecasting system, no future information is used for the estimation of seizure likelihood [Mormann_2007]. We thus repeated our analysis by ensuring that no future information is used in the test dataset. Specifically, we z-normalized the features of the test patient using the mean and standard deviation of the feature vectors that corresponded to the first two hours of iEEG recordings. We then excluded these two hours from the analysis and the algorithm was evaluated on the remaining dataset. This approach required to have at least five hours of iEEG recordings prior to the seizure occurrence (i.e., two hours of interictal data for the normalization and three hours of preictal data) and it was feasible to be tested in five patients (KCL1, KCL2, KCL3, KCL4, KCL7).

When we considered the time interval from 4000ms to 4980ms after the first stimulus (Figure S3A) the average IoC was 0.59. When we analysed the -1000ms to -20ms window prior to the first stimulus (Figure S3B), the average IoC was 0.34 across all patients. The seizure likelihoods as well as the forecasting metrics are illustrated in Figure S3.

### Impact of number of electrodes on forecasting performance

We also examined the effect of the number of analysed electrodes on the forecasts. We thus computed the forecasts using one iEEG channel (channel with most prominent response across stimulation procedure), five channels (top five channels with the most obvious responses) and all iEEG cannels in each patient. We performed this analysis using the features computed from the window 4000ms to 4980ms after the first stimulus (Figure 4A) and features computed from the window -1000ms to -20ms prior to the administration of the first stimulus (Figure 4B). We observed that the best performance was achieved when we considered five iEEG channels. In addition, when we analysed one or five iEEG channels the forecasts that were estimated from the window 4000ms to 4980ms after the first stimulus (Figure 4C; average IoC for one and five channels: 0.34; 0.74 respectively) outperformed the forecasts that were computed from the window -1000ms to -20ms prior to the administration of the first stimulus (Figure 4D; average IoC for one and five channels: 0.1; 0.54 respectively). When we considered all channels in the analysis the forecasting performance was the same (average IoC: 0.34) between the post-stimulation and pre-stimulation. The exact values of the forecasting performance are given in Tables S1-S6.

**Figure 4:**
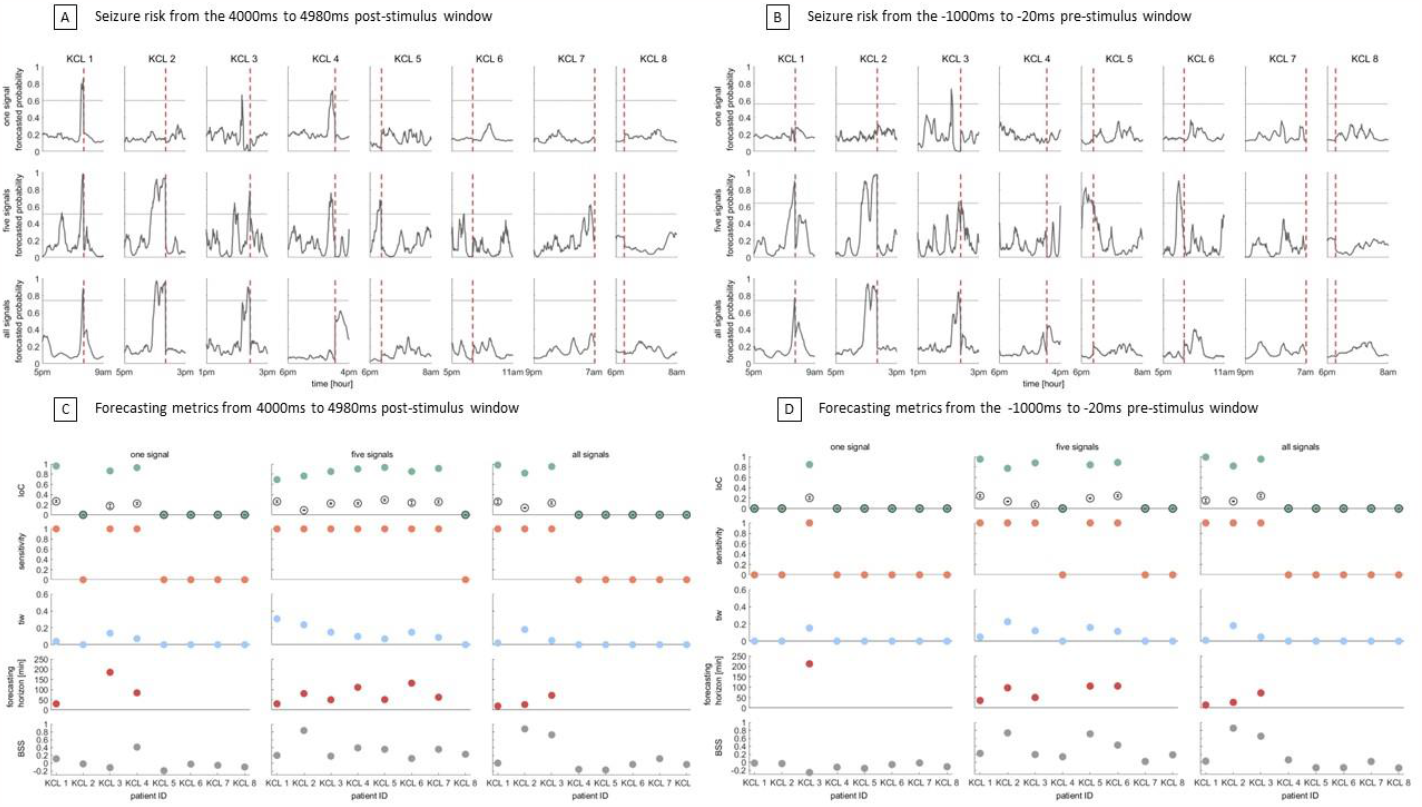
Seizure likelihood and forecasting metrics computed from the window 4000ms to 4980ms after the first stimulus (A, C) the -1000ms to -20ms window prior to the first stimulus (B, D), using one, five and all iEEG electrodes. The vertical dashed line denotes the seizure onset. After grid search (see Methods) the probability threshold (horizontal line) and seizure occurrence period were set to 0.6; 210min (A, one signal), 0.51; 150min (A, five signals), 0.74; 150min (A, all signals) and 0.56; 240min (B, one signal), 0.64; 120min (B, five signals), 0.74; 150min (B, all signals) respectively. Grey unfilled circles in panels C and D denote the average IoC obtained from the shuffled forecasts across 100 runs, whilst error bars denote the standard error. Note that negative IoC values from the shuffled forecasts were set to zero prior to averaging.

## Discussion

In this study we analysed a cohort of eight people with treatment-resistant focal epilepsy who underwent intermittent repeated electrical stimulation for approximately one day. We computed quantitative features from various iEEG windows relative to the stimuli, and showed that, using the window 4000ms to 4980ms after the first stimulus, seizures can be forecast above chance levels in seven out of eight patients. In addition, in the analysed cohort of patients we found that probing the brain with electrical stimulation is more informative for seizure forecasting compared to passive monitoring without stimulation.

Short-term post stimulus responses to SPES are widely used in presurgical evaluation for cortical mapping [Lacruz_2007, Keller_2014, Matsumoto_2017]. Specifically, early responses to SPES that occur within 100ms after stimulation are used to map functional connectivity of the motor cortex and language areas. In addition, late responses to SPES that occur from 100ms to 1sec after stimulation are used to identify the epileptogenic tissue [Valentin_2002, Valentin_2005]. In this study we found that short-term post-stimulus responses that consider iEEG intervals up to 100ms after stimulation were not informative for seizure forecasting (Figure S2A and S2B). When we considered longer post-stimulus intervals that encompass delayed responses (i.e., 1000ms after each stimulus), we were able to achieve seizure forecasting in two out of eight patients (Figure S2C and S2D). However, this was only possible when we analyzed the cortical responses from the second stimulus. This finding may indicate that each stimulus carries different information and therefore it may be more informative if stimuli are analyzed separately [Cornblath_2023]. In addition, those findings might indicate that the second stimulus is more informative compared to the first one due to the presence of a possible prolonged cortical excitability effect from the first stimulus (note that the time gap between the first and second stimuli was 5sec).

When we analysed the long-term post-stimulus responses of the first stimulus (i.e., 4000ms to 4980ms after the stimulus) we found that, in seven out of eight patients, seizures could be forecast above chance levels (Figure 3A and 3B). Note that for patient KCL8, in whom forecast above chance was not achieved, there were only 84min of iEEG available prior to seizure onset and therefore the poor performance might be due to the limited data. In addition, we demonstrated that seizure forecasting using passive unstimulated iEEG was successful in five out of eight patients (Figure 3C and 3D). When we mimicked a real-world seizure forecasting system, we also found that active perturbation of the cortex and measuring its response is more informative for seizure forecasting compared to passive monitoring (Figure S3). These findings are in line with previous studies on theoretical models. Specifically, a computational model of TLE demonstrated that changes in excitability that precede epileptic seizures may be more informative for seizure anticipation compared to passive monitoring [Suffczynski_2008]. Moreover, a theoretical model using a probing stimulus can extract information from the EEG for seizure anticipation [O’Sullivan-Greene_2009]. In addition, other theoretical models demonstrated that seizure anticipation may be feasible by applying small perturbation in the cortical dynamics [Kalitzin_2010].

We found that the best seizure forecasting performance was achieved when we considered five iEEG electrodes. This finding holds for both post and pre-stimulus iEEG intervals (Figure 4). The channels that were selected for the constant stimulation belong to the suspected seizure focus. In addition, the five electrodes that we considered in the analysis were those that manifested the most obvious responses during the stimulation, and hence it is very likely that those electrodes are part of the seizure onset network. Considering one channel in the analysis might not be enough to capture all the changes in cortical excitability, whilst analysing all electrodes might add redundant information. Future studies are needed to identify the optimal number and placement of electrodes for optimal seizure forecasting performance.

The seizure forecasting algorithm deployed a logistic regression model which is considered one of the simplest classifiers to obtain probability forecasts. The selection of this model in combination with the leave-one-patient out cross validation approach reduced the possibility of model overfitting. The main quantitative features that were employed in the seizure forecasting algorithm were the variance and autocorrelation. Previous studies [Mormann_2005, Maturana_2020] that analysed passively collected iEEG recordings showed that those features are informative for seizure forecasting. In addition, it has been shown that in the phenomenon of “critical slowing down” in which dynamical systems take longer time to return to equilibrium after perturbations there is an increase in the signal variance and autocorrelation [Scheffer_2009]. Furthermore, it has been shown in theoretical models that the variance is a metric that captures the energy between signals [Laiou_2017]. Additional features that were employed in the logistic regression classifier were the cumulative average of variance and autocorrelation to allow the model to consider the history and the evolution of those features. Future studies with larger cohorts of patients and longer recordings should deploy more advanced machine and deep learning approaches to optimize the seizure forecasting performance.

To the best of our knowledge this is the first study to show that data from a time-window 4-5 sec after stimulation may be informative for seizure forecasting. Due to the limited amount of available iEEG data, we were not able to verify this finding using the second stimulus, nor explore whether the optimal long-term post-stimulation window might be even longer after the stimulus; these considerations await future studies.

Although the findings of this study show promise for seizure forecasting, they have to be interpreted with caution. First, the analysed cohort was small, and no definitive conclusions can be made. In addition, the absence of multiday recordings did not allow us to investigate the presence of circadian or multidien cycles on cortical excitability as well as to include time-matched seizure surrogate data [Andrzejak_2003]. Moreover, we analysed only one seizure per patient. Future studies with longer data should investigate whether the same findings hold for patients who manifest multiple seizures. In such cases the seizure forecasting parameters could be also optimized for each patient individually to enhance the seizure forecasting performance.

In conclusion, this study adds to previous experimental and theoretical studies [Kalitzin_2002, Kalitzin_2005, Suffczynski_2008, Kalitzin_2010, Freestone_2011] which showed that seizure forecasting may be possible by probing the brain with electrical stimulation. Additionally, this work demonstrates that late post-stimulus iEEG intervals may be more informative for seizure forecasting compared to iEEG intervals that correspond to passive monitoring of the brain. These findings may not only aid in the development of seizure forecasting algorithms but also in the design of novel implantable devices that deliver electrical stimulation to control seizures. The use of neuromodulation devices will be expanded in the near future [Denison_2022] and we hope that this work will motivate further research into uncovering the use of cortical electrical stimulation as a tool for seizure forecasting.

## Data availability statement

The analysed data are available from the corresponding authors upon reasonable request.

## Author contribution

Formal analysis: P.L., Methodology: P.L., Z.K., M.D., Data curation: A.V., S.J., J.S.W., P.V., Writing-original draft: P.L., M.P.R., Writing-review and editing: P.L., Z.K., A.V., S.J., M.C., P.V., R.D., A.S.-B., M.D., T.J.D, J.S.W., M.P.R., Funding acquisition: P.L., J.S.W., M.P.R, Supervision: M.P.R

## Acknowledgements

We would like to acknowledge Drs Amir Eftekhar, Richard Selway, Gonzalo Alarcon and Andrea Biondi for assistance with data collection and useful discussions.

