## Supplementary material for "Seizure forecasting by tracking cortical response to electrical stimulation"

Figures

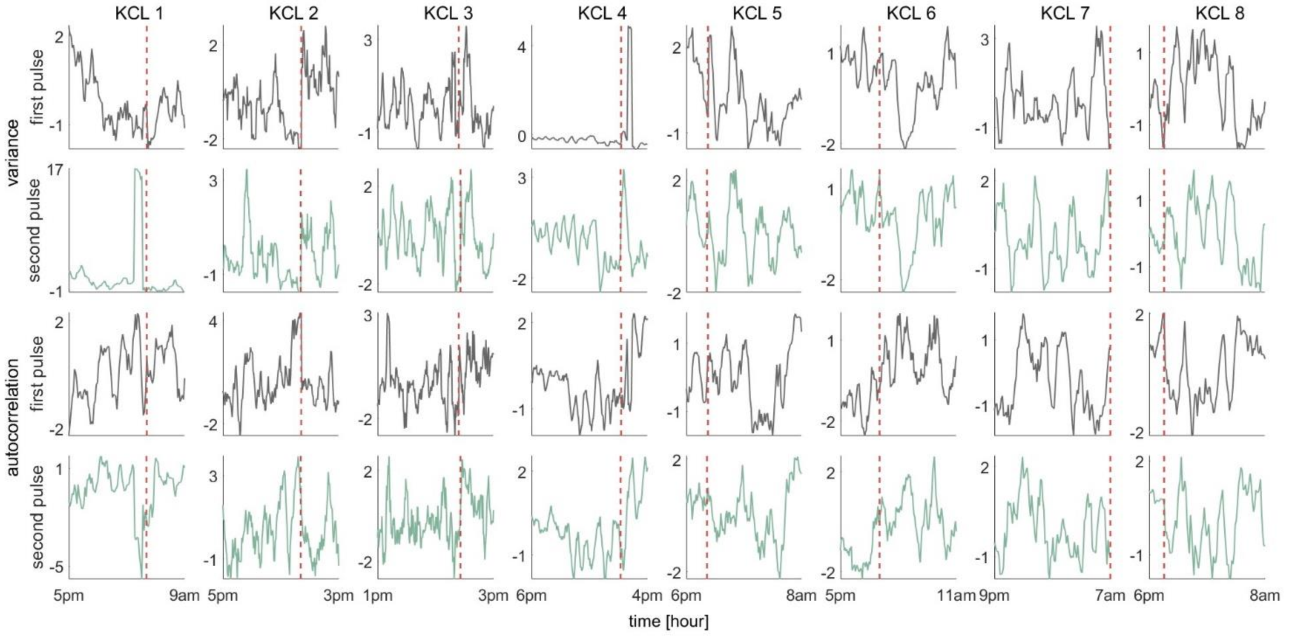


Figure S1: Temporal evolution of the features from the window 20ms to 100ms after the first stimulus (grey) and after the second stimulus (green). The vertical dashed line denotes the seizure onset.

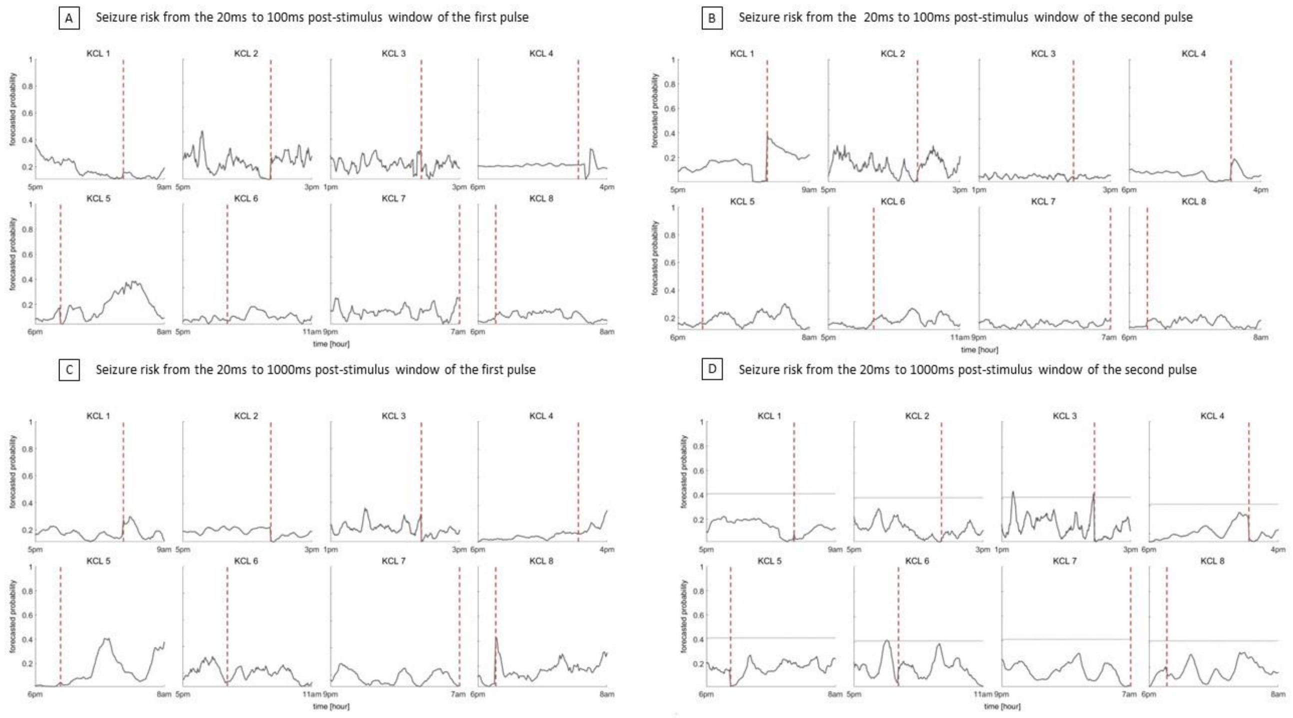


Figure S2: Seizure likelihood computed from the windows 20ms to 100ms and 20ms to 1000ms after the first (A, C) and second stimulus (B, D). The vertical dashed line denotes the seizure onset. All patients in panels A, B and C have zero Improvement over chance. The grid search (see Methods) in patients of panel D set the probability threshold (horizontal line) to 0.41 and the seizure occurrence period to 90min. Forecasting metrics in panel D: KCL3 (sensitivity:1, tiw: 0.08, IoC: 0.92, forecasting horizon: 17.7min, BSS: 0.03) and KCL 4 (sensitivity:1, tiw: 0.09, IoC: 0.9, forecasting horizon: 88.9 min, BSS: 0.07) respectively.

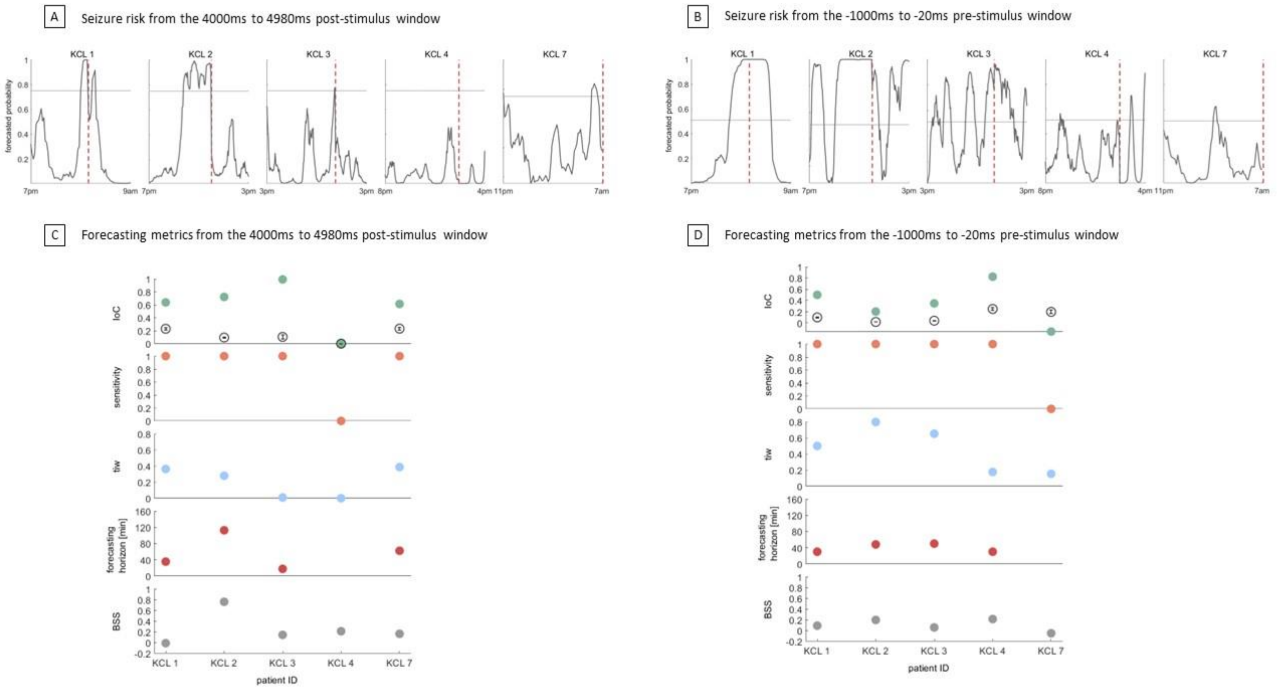


Figure S3: Seizure likelihood and forecasting metrics computed from the window 4000ms to 4980ms after the first stimulus (A, C) and the -1000ms to -20ms window prior to the first stimulus (B, D). Note that during the leave-one-subject-out approach no future information was used in the test patient. The vertical dashed line denotes the seizure onset. After grid search (see Methods) the probability threshold (horizontal line) and seizure occurrence period were set to 0.75; 120min (A) and 0.51; 60min(B) respectively. Gray unfilled circles in panels C and D denote the average IoC obtained from the shuffled forecasts across 100 runs, whilst error bars denote the standard error. Note that negative IoC values from the shuffled forecasts were set to zero prior to averaging.

### Tables

| | KCL 1 | KCL 2 | KCL 3 | KCL 4 | KCL 5 | KCL 6 | KCL 7 | KCL 8 |
|---|---|---|---|---|---|---|---|---|
| Sensitivity | 1 | 1 | 1 | 1 | 1 | 1 | 1 | 0 |
| IoC | 0.69 | 0.76 | 0.85 | 0.9 | 0.93 | 0.85 | 0.91 | 0 |
| Mean (IoC); standard error | 0.26; 0.024 | 0.09; 0.0059 | 0.22; 0.018 | 0.23; 0.019 | 0.29; 0.017 | 0.24; 0.037 | 0.26; 0.023 | 0; 0 |
| Tiw | 0.31 | 0.24 | 0.15 | 0.097 | 0.068 | 0.15 | 0.087 | 0 |
| Forecasting horizon (min) | 30 | 81 | 50 | 111 | 51 | 132 | 62 | N/A |
| BSS | 0.2; 0.0046 | 0.83; 0.0012 | 0.18; 0.0039 | 0.39; 0.003 | 0.35; 0.0038 | 0.12; 0.0041 | 0.35; 0.0034 | 0.23; 0.0036 |

Table S1: Forecasting values when we considered in the analysis the 4000ms to 4980ms window after the first stimulus using five signals.

| | KCL 1 | KCL 2 | KCL 3 | KCL 4 | KCL 5 | KCL 6 | KCL 7 | KCL 8 |
|---|---|---|---|---|---|---|---|---|
| Sensitivity | 1 | 1 | 1 | 0 | 1 | 1 | 0 | 0 |
| IoC | 0.95 | 0.77 | 0.88 | 0 | 0.84 | 0.89 | 0 | 0 |
| Mean (IoC); standard error | 0.24; 0.027 | 0.14; 0.0072 | 0.078; 0.025 | 0; 0 | 0.2; 0.012 | 0.24; 0.023 | 0; 0 | 0; 0 |
| Tiw | 0.049 | 0.23 | 0.12 | 0 | 0.16 | 0.11 | 0 | 0 |
| Forecasting horizon (min) | 36 | 97 | 50 | N/A | 104 | 105 | N/A | N/A |

| | | | | | | | | |
|---|---|---|---|---|---|---|---|---|
| BSS | 0.22; 0.0059 | 0.74; 0.0019 | 0.19; 0.0038 | 0.14; 0.0029 | 0.71; 0.0024 | 0.43; 0.004 | 0.021; 0.0035 | 0.19; 0.003 |

Table S2: Forecasting values when we considered in the analysis the -1000ms to -20ms window before the first stimulus using five signals.

| | KCL 1 | KCL 2 | KCL 3 | KCL 4 | KCL 5 | KCL 6 | KCL 7 | KCL 8 |
|---|---|---|---|---|---|---|---|---|
| Sensitivity | 1 | 0 | 1 | 1 | 0 | 0 | 0 | 0 |
| IoC | 0.96 | 0 | 0.86 | 0.93 | 0 | 0 | 0 | 0 |
| Mean (IoC); standard error | 0.27; 0.024 | 0; 0 | 0.17; 0.036 | 0.22; 0.022 | 0; 0 | 0; 0 | 0; 0 | 0; 0 |
| Tiw | 0.04 | 0 | 0.14 | 0.072 | 0 | 0 | 0 | 0 |
| Forecasting horizon | 30 | N/A | 190 | 84 | N/A | N/A | N/A | N/A |
| BSS | 0.11; 0.0045 | -0.019; 0.0016 | -0.11; 0.0035 | 0.41; 0.0021 | -0.19; 0.0034 | -0.024; 0.0021 | -0.056; 0.0015 | -0.099; 0.0027 |

Table S3: Forecasting values when we considered in the analysis the 4000ms to 4980ms window after the first stimulus using one signal.

| | KCL 1 | KCL 2 | KCL 3 | KCL 4 | KCL 5 | KCL 6 | KCL 7 | KCL 8 |
|---|---|---|---|---|---|---|---|---|
| Sensitivity | 1 | 1 | 1 | 0 | 0 | 0 | 0 | 0 |
| IoC | 0.98 | 0.82 | 0.95 | 0 | 0 | 0 | 0 | 0 |
| Mean (IoC); standard error | 0.26; 0.034 | 0.14; 0.0079 | 0.23; 0.023 | 0; 0 | 0; 0 | 0; 0 | 0; 0 | 0; 0 |
| Tiw | 0.02 | 0.18 | 0.049 | 0 | 0 | 0 | 0 | 0 |
| Forecasting horizon (min) | 19 | 27 | 72 | N/A | N/A | N/A | N/A | N/A |
| BSS | 0.0027; 0.0055 | 0.88; 0.0009 | 0.72; 0.0012 | -0.16; 0.0069 | -0.17; 0.0032 | -0.032; 0.0027 | 0.11; 0.0025 | -0.034; 0.0028 |

Tale S4: Forecasting values when we considered in the analysis the 4000ms to 4980ms window after the first stimulus using all signals.

| | KCL 1 | KCL 2 | KCL 3 | KCL 4 | KCL 5 | KCL 6 | KCL 7 | KCL 8 |
|---|---|---|---|---|---|---|---|---|
| Sensitivity | 0 | 0 | 1 | 0 | 0 | 0 | 0 | 0 |
| IoC | 0 | 0 | 0.85 | 0 | 0 | 0 | 0 | 0 |
| Mean (IoC); standard error | 0; 0 | 0; 0 | 0.21; 0.028 | 0 | 0 | 0 | 0 | 0 |
| Tiw | 0 | 0 | 0.15 | 0 | 0 | 0 | 0 | 0 |
| Forecasting horizon | N/A | N/A | 212 | N/A | N/A | N/A | N/A | N/A |
| BSS | -0.02; 0.0015 | -0.032; 0.002 | -0.26; 0.0052 | -0.12; 0.0024 | -0.15; 0.0032 | -0.057; 0.0022 | -0.015; 0.0023 | -0.11; 0.0038 |

Table S5: Forecasting values when we considered in the analysis the -1000ms to -20ms window before the first stimulus using one signal.

| | KCL 1 | KCL 2 | KCL 3 | KCL 4 | KCL 5 | KCL 6 | KCL 7 | KCL 8 |
|---|---|---|---|---|---|---|---|---|
| Sensitivity | 1 | 1 | 1 | 0 | 0 | 0 | 0 | 0 |
| IoC | 0.99 | 0.82 | 0.95 | 0 | 0 | 0 | 0 | 0 |
| Mean (IoC); standard error | 0.16; 0.034 | 0.14; 0.0095 | 0.24; 0.03 | 0; 0 | 0; 0 | 0; 0 | 0; 0 | 0; 0 |
| Tiw | 0.0098 | 0.18 | 0.048 | 0 | 0 | 0 | 0 | 0 |
| Forecasting horizon (min) | 14 | 27 | 72 | N/A | N/A | N/A | N/A | N/A |
| BSS | 0.028; 0.0043 | 0.85; 0.0009 | 0.65; 0.0017 | 0.061; 0.0035 | -0.14; 0.0024 | -0.13; 0.004 | 0.02; 0.0021 | -0.14; 0.0034 |

Table S6: Forecasting values when we considered in the analysis the -1000ms to -20ms window before the first stimulus using all signals.
